# Improving UK data on avoidable perinatal brain injury: consultations and co-creation of recommendations

**DOI:** 10.64898/2026.05.01.26352206

**Authors:** Jan W. van der Scheer, Kirstin Webster, Muhammed Ally Hussein Wahedally, Jane O’Hara, Tim Draycott, Andrew Demitri, Rachna Bahl, ABC Data Advisory Group, ABC PPI Group, Mary Dixon-Woods

**Affiliations:** THIS Institute (The Healthcare Improvement Studies Institute), University of Cambridge, Strangeways Research Laboratory, Cambridge CB1 8RN, UK; Royal College of Obstetricians & Gynaecologists, 10-18 Union Street, London, SE1 1SZ, UK; Department of Population Health Sciences, University of Leicester, Leicester, LE1 7RH, UK; University of Bristol, Beacon House, Queens Road, Bristol, BS8 1QU, UK; University Hospitals Bristol and Weston, Bristol, BS2 8EG, UK

## Abstract

**Objective:** Avoidable perinatal brain injury, including preventable hypoxic⍰ischaemic encephalopathy (HIE), remains a major global challenge for maternity care. In the UK, progress is hindered by weaknesses in infrastructure of routinely collected perinatal data. We sought to develop a strategy to support improvement in data relating to potentially avoidable HIE.

**Design:** Online survey of UK-based professionals, structured discussions with a multidisciplinary advisory group, and three workshops with patient/family representatives.

**Setting:** United Kingdom.

**Main outcome measures:** Co-created recommendations for improving the consistency, comprehensiveness, linkage, quality, and practical use of routinely collected UK maternity and neonatal data related to potentially avoidable HIE.

**Results:** Between 85–98% of survey participants (*N*=411) rated most of the data items proposed as characterising avoidable brain injury as important. They also identified data gaps, particularly for intrapartum risk factors. Participants wanted accessible, unit⍰level feedback and capability⍰building for interpreting data, while cautioning against sharing data with families without context. The advisory group (*n*=35) and patient/family representatives (*n*=9) converged on 15 recommendations covering: definitions; a core item catalogue and shared data dictionary; electronic patient record interoperability; workflow⍰integrated data capture; secure individual⍰level linkage with longer⍰term follow⍰up; strengthened audit and feedback; and improved capture and sharing of data relevant to families.

**Conclusions:** The recommendations offer a roadmap for developing an integrated data source that builds on existing datasets of routinely collected maternity and neonatal data. Even a basic version of this data source would already help promote actionable use of data and guide investments in digital infrastructure that enables continuous improvement cycles.

**Key messages:** *What is already known on this topic:*

- Efforts to reduce avoidable perinatal brain injury in the UK are hindered by fragmented maternity and neonatal data systems, including inconsistent definitions, limited linkage across datasets, duplication in data entry practices, and incomplete coverage of key data items.

*What this study adds:*

- This study presents strategic recommendations for improving UK data on potentially avoidable perinatal brain injury, collaboratively developed with maternity and neonatal professionals, data specialists, and patient and family representatives.
- The 15 recommendations set out ways to strengthen definitions, comprehensiveness, standardisation, linkage, quality, and practical use of routinely collected data relevant to avoidable perinatal brain injury.

*How this study might affect research, practice or policy:*

- The set of recommendations offers a roadmap for developing an integrated data source that builds on existing datasets of routinely collected maternity and neonatal data.
- Even a basic version of an integrated data source would already help promote meaningful and actionable use of data for prevention of perinatal brain injury, and guide future investment in digital infrastructure that enables continuous improvement cycles.

## Introduction

Hypoxic-ischaemic encephalopathy (HIE) is a major cause of avoidable perinatal brain injury.^1^ HIE is one of the leading causes of neurodisability and mortality in near-term and term babies in high-income countries such as the UK, with estimates of average incidence ranging from 1.5 to 2.0 per 1000 live births.^2–7^ Mild, moderate and severe forms of HIE and other potentially avoidable brain injury occurring around the time of birth can have devastating and long-term consequences for babies and families,^2,3,8–10^ generate substantial lifetime costs for health and care services,^11^ and can lead to clinical negligence settlements exceeding £20m per avoidable case of cerebral palsy in the UK.^12,13^

The causes of avoidable HIE-related brain injury are multi-factorial and complex.^1,12–16^ To address its causes and reduce incidence, professionals need insights from high-quality, standardised, and meaningful routinely collected data.^17^ Such data enable the monitoring of incidence, identification of risk factors, understanding of variation, quality assurance, and ultimately better care in maternity and neonatal services.^4,6,18–21^ The maternity and neonatal data landscape in the UK is extensive but fragmented: brain injury-relevant data are captured across multiple national sources,^17,22^ with inconsistencies in definitions and recording practices,^17,22,23^ and limited linkage across maternity and neonatal datasets.^17,18,22,23^ Similar challenges have been recognised internationally,^17,21,24–26^ underscoring the need for coordinated strategies to strengthen perinatal data systems.

Addressing these data challenges in maternity and neonatal care requires a strategy that combines technical solutions – such as standardising definitions and improving interoperability of electronic patient record (EPR) platforms^22,23^ – with attention to socio-technical factors, including clinical workflows, data entry practices, and how insights from data might be interpreted and actioned by professionals and families.^22^ A co-created approach – defined as meaningful involvement of professionals, families, and other relevant specialists across all development stages, from identifying problems to exploring potential solutions – can help ensure that future data strategies are responsive to real-world needs and feasible in practice.^22,27–29^

We aimed to co-create a strategy for developing an integrated UK data source on potentially avoidable brain injury around the time of birth, with a focus on HIE. Building on our earlier data dictionary review and multidisciplinary consultation,^22^ our objectives were to:

i. undertake a survey of UK-based maternity and neonatal professionals to explore views on the design and use of an integrated data source, and
ii. iteratively consult a multidisciplinary group and patient/family representatives to agree on recommendations for improving data related to avoidable HIE-related brain injury.

## Methods

We undertook three stages of work to co-create the recommendations (**Table 1**). Stage 1 (2022) comprised a review of data dictionaries of four UK maternal and neonatal data sources, to identify data limitations and inform questions/topics for subsequent consultations. Stage 2 (2022-2025) involved consultations with professionals and patient/family representatives, with each consultation informing the next. In Stage 3 (2025-2026), findings of the review and consultations informed a draft set of recommendations, which was then iteratively shaped and agreed in collaboration with a multidisciplinary advisory group and patient/family representatives.

**Table 1.** Activities and key findings underpinning the co-creation of recommendations for improving data related to avoidable HIE-related brain injury around the time of birth.

| Stage | Study design | Participants | Data collection | Key findings |
| --- | --- | --- | --- | --- |
| **Stage 1** | Data dictionary review (2022)<br>Further detail: van der Scheer et al<sup>22</sup> | Not applicable. | Review of data dictionaries of four main UK maternal/neonatal data sources (MSDS, HES APC, NNRD, NMPA) to: (i) identify which of their data items are relevant to clinical indicators or risk factors of avoidable HIE-related brain injury, and (ii) assess homogeneity of definitions of similar data items across the data sources. | The review showed many limitations of current data sources: none of them on their own captures the range of items relevant to injury, key intrapartum risk factors are not consistently captured, operational definitions of similar data items differ across data sources, and a mechanism for ongoing linkage of data across data sources is absent. |
| **Stage 2a** | Multidisciplinary interview consultation (2022)<br>Further detail: van der Scheer et al<sup>22</sup> | Purposively invited UK-based multidisciplinary group (N=27) including individuals with leadership roles or expertise spanning neonatal care, maternity care, statistics, and clinical negligence. | The individual interviews explored views on relevant data items and definitions of avoidable HIE-related brain injury as well as current challenges in capturing and using data. | The consultation identified the need for standardising the definition of avoidable brain injury around the time of birth, resolving inconsistencies in capturing data, improving linkage of data across existing data sources, and co-creating a strategy for meaningful use of data. |
| **Stage 2b** | Survey of UK-based maternity and neonatal professionals (2024-2025) | Convenience sample of 411 professionals from all UK regions, including roughly 70% maternity (consultant and trainee obstetricians, senior and junior midwives, and midwives with digital roles) and 30% neonatal (consultant and trainee neonatologists, advanced nurse practitioners, and allied health professionals). | The online survey used the Thiscovery (https://thiscovery.org/) platform to gather views on: (i) defining avoidable brain injury around the time of birth, (ii) data items for a future integrated data source, and (iii) using routinely collected data to support improvements in care. Further detail: Supplement 1. | See: Results section in this paper, including Box 1, Table 2, Table 3, and Supplement 3. |
| **Stage 2c** | Online discussions with multidisciplinary advisory group (2025) | Purposively invited UK-based multi-disciplinary advisory group, including 35 individuals with leadership roles or expertise | The group was consulted on the Thiscovery platform through structured briefings, short surveys, and optional forum discussions to: (i) inform defining | See: Results section in this paper, including Supplement 2 and Supplement 3. |
|  |  | spanning obstetrics, midwifery, neonatology, paediatrics, anaesthetics, clinical audit, epidemiology, statistics, safety investigations, and medical litigation. | avoidable brain injury in the perinatal context, (ii) assess and recommend data items for an integrated source, and (ii) review draft recommendations that drew on issues identified in the data dictionary review and interview consultation (see above).<sup>22</sup> Further detail: Supplement 2. |  |
| **Stage 2d** | Workshops with patient and family representatives (2025)<br>Further detail: Supplement 4 | Purposively invited UK-based group of women with experience of UK maternity services, including 9 maternity safety advocates and representatives from charities supporting women and families (particularly those affected by perinatal injury and those whose voices are under-represented). | Three online semi-structured workshop discussions, facilitated by a healthcare improvement specialist and a neonatal professional, to explore lived experience and views on: (i) what types of brain injury-related data are most meaningful to families, (ii) how data should be shared to support decision-making, and (iii) what good communication looks like in the context of sensitive information. Further detail: Supplement 4. | See: Results section in this paper, including Supplement 3 and Supplement 4 |
| **Stage 3** | Revising and agreeing recommendations (2025-2026)<br>Further detail: Supplement 5 | Multidisciplinary advisory group (N=35) and patient and family representatives (N=9) who participated in Stage 2. | After incorporating feedback on the draft recommendations (Supplement 2), the advisory group agreed (using an asynchronous consensus exercise conducted over email) on recommendations covering definitions, data alignment, data linkage, data quality, feasibility, and data-driven feedback for professionals. Summaries from patient/family workshops (Supplement 4) and draft recommendations on data sharing were shared with the patient/family representatives, with iterative email revisions leading to agreement on two family-facing recommendations | Agreed set of 15 recommendations around definitions, aligning data, linkage of data sources, data quality, feasibility of data capture, data-driven feedback mechanisms for professionals, and improving data and data sharing relevant to families (Table 4). |

Abbreviations: Maternity Services Data Set (MSDS), Hospital Episode Statistics for Admitted Patient Care (HES APC), the National Neonatal Research Database (NNRD), National Maternity and Perinatal Audit (NMPA).

## Results

**Table 1** presents high-level findings of the review (Stage 1) and multidisciplinary interviews (Stage 2a), which are reported in detail elsewhere.^22^ Below, we present the findings of the other consultations (Stages 2b, 2c and 2d in **Table 1**), followed by the co-created recommendations (Stage 3).

### Defining avoidable brain injury around the time of birth

Most survey participants (*n*=256, 62%) indicated that **HIE** best described the clinical condition relevant to monitoring incidence and risk factors of avoidable brain injury around the time of birth. Free⍰text responses emphasised that while HIE is a useful core diagnostic descriptor, any definition must acknowledge the challenges of attributing causality.

> *“[A]voidable is very subjective and difficult to define even after rigorous review. I would feel anxious that we use […] the word ‘avoidable’ too liberally.” (Consultant obstetrician)*

> *“In a detailed definition, it is essential to stress the timing, preventable factors (like substandard care), and outcomes. By considering circumstances like congenital anomalies or rare complications, it aims to differentiate between preventable and unavoidable injuries clearly, guiding enhancements in perinatal care to minimize avoidable harm.” (Senior midwife)*

Most advisory group members (*n*=31, 86%) also agreed that **signs and symptoms consistent with HIE** should form part of a definition to underpin an integrated data source. They noted that this focus is justified because HIE is the leading contributor to potentially avoidable brain injury in the UK, aligns with current UK guidelines for therapeutic hypothermia, and is relatively consistently identifiable in structured fields within existing datasets. The advisory group further agreed that the definition should focus on **infants at or near term (**≥**36 weeks)**, consistent with eligibility criteria for therapeutic hypothermia and reflecting the greater complexity of assessing avoidable causes of HIE-like signs in preterm infants (who are also already captured in neonatal data reporting). They also supported including **infants with mild HIE**, acknowledging that such cases may be under-diagnosed and – in contrast to moderate or severe HIE – are less reliably captured in neonatal reporting given that babies with signs of mild HIE are not always admitted to a neonatal unit.

### Data items for an integrated data source

The survey included a set of possible clinical indicators and risk factors that might characterise avoidable brain injury and would be relevant to include in an integrated data source. Over 80% of participants rated most items in this set as important or extremely important (85-98%, **Table 2**). Fewer viewed maternity characteristics as important (73%, **Table 2**), with free-text comments suggesting that such characteristics are relevant only when linked to pre⍰existing risk factors or when they may have an impact on care, with specific attention to the limitations of ethnicity⍰related data.

> *“When describing ethnicity, care should be taken to acknowledge limitations on data (e.g. groupings used such as “Asian”) and avoid any risk of poorly designed clinical decision making tools using ethnicity that could become “self fulfilling prophecies”” (Registrar neonatologist)*

**Table 2.**
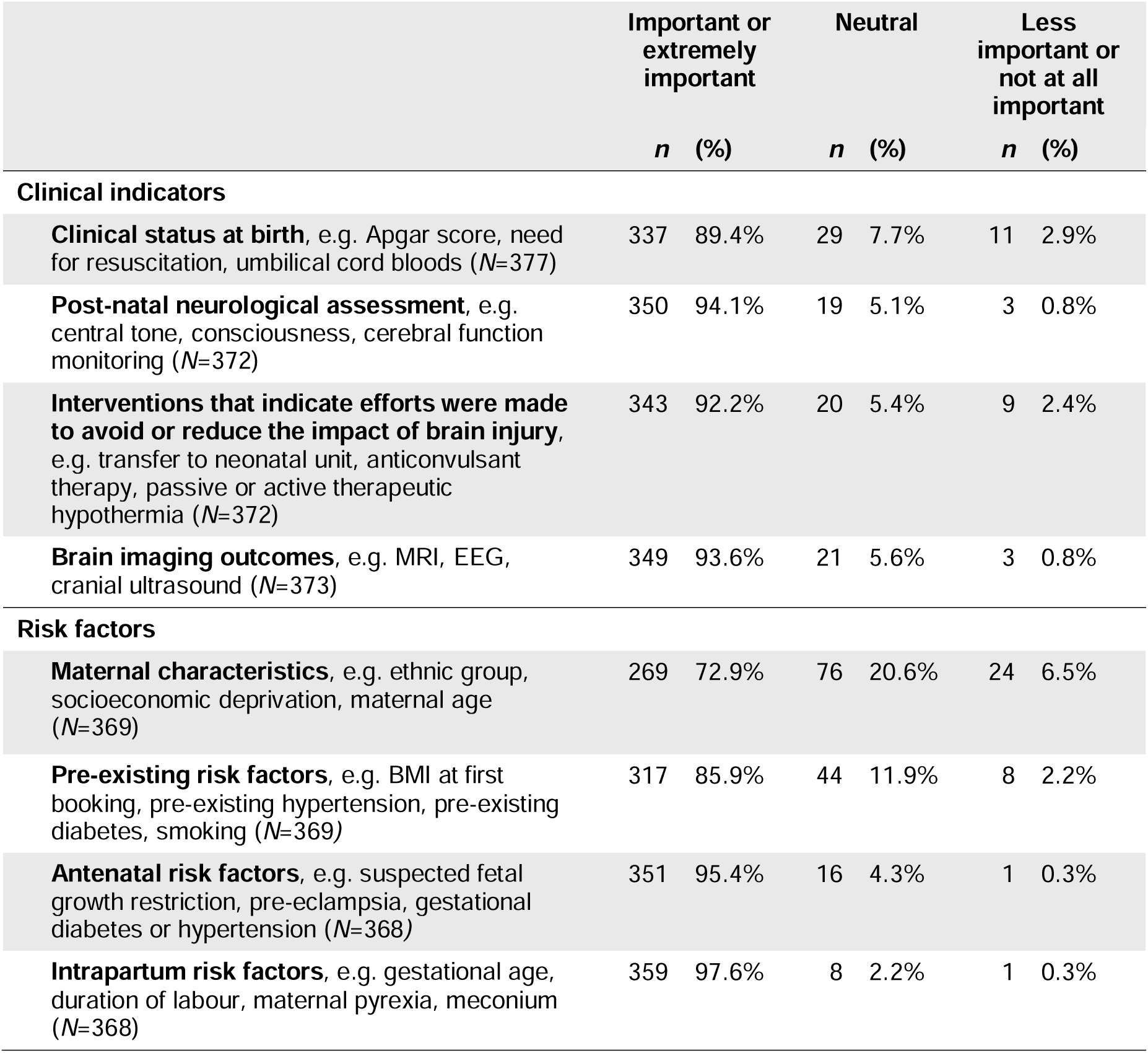
Survey participants’ ratings of the importance of clinical indicators and risk factors for understanding potentially avoidable brain injury around the time of birth. *N* indicates total number of responses.

Survey participants were also asked to identify which of a range of pre⍰specified data items – selected from our previous data dictionary review^22^ as items that may be missing from existing national datasets – would be important to include in an integrated data source. From the 367 participants who responded (**Supplement 3**), 80% or more selected the occurrence of placental abruption (n=329, 90%) or shoulder dystocia (n=327, 89%), the classification of intrapartum cardiotocography (n=309, 84%), and whether intermittent auscultation and/or cardiotocography was used (n=293, 80%). A subset of survey participants (*n*=34) and advisory group members (*n*=22) suggested additional items they considered important to include in an integrated data source (**Supplement 3**), including:

- antenatal risk factors (e.g. concerns about antenatal fetal movements, care outside clinical guidance),
- intrapartum risk factors (e.g. cardiotocography classification accuracy, duration of fetal heart rate abnormality, occurrence of cord prolapse, fetal haemorrhage, impacted fetal head).
- intervention processes (e.g. timing of cord clamping, time-to-initiation of therapeutic hypothermia, therapeutic temperature maintenance during transfer), and
- post-natal assessments (e.g. placental histology, neonatal withdrawal scores).

Patient/family representatives (n=9) noted that several aspects of care important to families appear to be absent from current national datasets. Key examples included whether those in labour received continuous one⍰to⍰one support, whether they felt physically and emotionally safe, whether families’ concerns were acknowledged and acted upon during antenatal or intrapartum care, and whether antenatal diagnostic opportunities were missed or delayed (**Supplement 3**). Representatives explained that developing structured items in these areas could help identify service⍰level patterns, strengthen trust between service users and providers, and ensure data better reflects what contributes to feeling safe, heard, and supported.

### Using routinely collected data to support improvements in care

Survey participants showed a strong appetite for getting better insights into summarised data relating specifically to their own maternity/neonatal unit and training in using data to improve their practice (**Table 3**). They provided a range of suggestions how to improve this (**Box 1**).

**Table 3.** Survey participants’ ratings of agreement with statements on sharing and using data to improve practice. *N* indicates total number of responses.

| Statements | Agree or strongly agree |  | Neutral |  | Disagree or strongly disagree |  |
| --- | --- | --- | --- | --- | --- | --- |
|  | *n* | (%) | *n* | (%) | *n* | (%) |
| Maternity/neonatal professionals should get better insights into summarised data on avoidable brain injury in their own unit (*N*=338) | 311 | 92.0% | 21 | 6.2% | 6 | 1.8% |
| Maternity/neonatal professionals are more likely to improve their practice if data on avoidable brain injury in their own unit were presented in an accessible and meaningful way (*N*=337) | 310 | 92.0% | 23 | 6.8% | 4 | 1.2% |
| Maternity/neonatal professionals need further training on how to improve their practice using data on avoidable brain injury of their own unit (*N*=338) | 265 | 78.4% | 63 | 18.6% | 10 | 3.0% |
| It's important for all units to routinely monitor and review their own data on avoidable brain injury (*N*=336) | 326 | 97.0% | 10 | 3.0% | 0 | 0.0% |
| Parents and people who are planning to be parents should have access to summarised data on avoidable brain injury in the unit they're attending (*N*=336) | 192 | 57.1% | 105 | 31.3% | 39 | 11.6% |

#### Box 1. Considerations and suggestions for developing and using an integrated data source

**Prioritising capturing and sharing relevant data**

> *“It seems that availability of HIE data is not recorded as a separate data set in my trust. We need to have this, so we can see rises and reductions over time. I know the numbers may be small compared to other types of data, but it would still be useful to understand, and teach this in training.” (Senior midwife)*

> *“We need large scale collection of good quality data but this should not be used to form a half baked league table. It should instead be used to inform practice, address inequality and support improvements in care” (Consultant neonatologist)*

**Improving data definitions in current datasets and integrating data capture to mitigate the “burden” on frontline staff**

> *“The flow of maternity data is truly shocking - I have enormous concerns about how poorly used maternity data is, which I attribute significantly to the structure of MSDS [Maternity Services Data Set]. The lack of development of basic definitions and dataflows has clearly done much to impede learning in the NHS.” (Consultant neonatologist)*

> *“[…] Having a separate database to enter data into however is always an extra burden of work for people on the busy shop floor. So if this could just be incorporated into [name of EPR supplier] for example, where midwives could put their data and neonates can put their side, this would likely reduce paperwork needed.” (Neonatal registrar)*

> *“Please make use of existing systems. I spend a ridiculous amount of time inputting data into multiple digital systems which don’t communicate with each other. It must be better to make better use of the digital systems we already use.” (Midwife)*

**Contextualising data based on the unit size and population the unit serves**

> *“This data needs to be contextualised and perhaps for comparison for other similar units/area of demographics. Also in context with other outcomes and markers of safe/quality care.” (Registered nurse/Midwife Qualified in Specialty)*

> *“Difficult to share data with parents to be as comparison between units can be misleading due to level of NICU that unit has, whether tertiary unit or not and other factors.” (Senior midwife)*

> *“[N]ational data should help to understand the trends rather than a league table.” (Consultant obstetrician)*

**Enhancing how units and individuals can learn from data**

> *“Data should guide the teams to interrogate and manage all risks relating to perinatal optimisation not only during the peripartum period but also before this period eg communications around placental issues and how this is acted on in subsequent pregnancies. I think it would be more beneficial if human factor experts are involved in the planning of this data interrogation and data interrogation is supported by guidance on what system elements will influence this data as most clinical people are not necessarily trained in systems thinking.” (Consultant neonatologist)*

> *“Avoid creating a punitive culture around data on avoidable brain injuries. Instead, focus on learning and improvement through case reviews, audits, and staff support. For example, real-time feedback mechanisms can make data more relatable and impactful for professionals. Training programs are needed to enhance healthcare professionals’ ability to interpret and use data effectively. Allocating resources for better electronic patient record systems and data analysis tools ensures higher accuracy and usability. Beyond immediate injuries, ongoing data collection on long-term developmental outcomes e.g cognitive or motor delays- is crucial. Understanding the broader impact of brain injuries informs better prevention and care pathways.” (Senior midwife)*

> *“Maybe better for units to buddy up and review data of ‘buddy’ against national standards? We don’t look at our own data with objectivity” (Job role not reported)*

**Improving training for clinicians around capturing and learning from data**

> *“Training clinicians in the importance of high quality data collection (real training where resulting in understanding and improved practice - not one slide or an IG [information governance] module!), [including] governance and assurance of data collection.” (Senior midwife)*

**Consider the risk of misinterpretation and undue stress if data is shared with families without sufficient context**

> *Parents having data out of context might be unhelpful for them to interpret and understand why a particular unit has higher rates. It can also be distressing and worrying if you live in an area where you don’t have choice of service due to geography. (Senior midwife)*

These findings – together with our earlier data dictionary review and multidisciplinary consultation,^22^ and literature relating to effective development, adoption and scale-up of data-driven feedback mechanisms (e.g.^20,30,31–33^) – informed a set of draft recommendations around developing and using an integrated data source to support improvements in care (**Supplement 2**). Twenty advisory group members reviewed these, with 75%–100% agreeing or strongly agreeing with the proposed recommendations, and provided feedback for improving the draft recommendations (**Supplement 2**).

### Sharing data with families

Survey views were mixed on sharing unit data on brain injury with families (e.g. 12% disagreed, **Table 3**), with free-text responses highlighting the risk of misinterpretation and undue stress (**Box 1**). This concern was echoed in workshops with patient/family representatives (**Supplement 4**), who emphasised that communication about injury-related data must be handled carefully, particularly when people may already feel anxious or overwhelmed.

> *“[…] you don’t want to present women with a long list of risk factors that will scare them, particularly when there isn’t always choice of an alternative hospital.” (Maternity service user)*

Patient/family representatives suggested antenatal discussion prompts and improved signposting to trusted information to support informed conversations between service users and care providers. They highlighted that data must be shared in a timely manner to support informed choice and active participation in care decisions.

> *“How does the data assist the parents’ ability to make informed choice, and / or be alert to concerns to promote their own / their child’s safety?” (Maternity service user)*

Consideration of health inequalities and equality, diversity and inclusion (EDI) was seen as key to any strategy around sharing data with families.

> *“Acknowledge that parents from ethnic minorities are more likely to experience adverse outcomes etcetera, are at greater risk etcetera, but have more access needs, [have] difficulty interpreting the data etcetera, [in turn] presenting challenges and ethical considerations.” (Maternity service user)*

It was also recognised that priorities for data vary. For example, people who have experienced trauma or bereavement may value understanding what has changed in response to past events rather than viewing statistical data only.

> *“Overall goal [should be] to contextualise adverse outcomes – women and people are often told “it’s a never event”, “it’s an isolated incident”. We need more narrative [that is] multifaceted.” (Maternity safety advocate)*

### Recommendations

Using an asynchronous consensus exercise conducted over email, the advisory group and the patient/family representatives shaped and agreed a set of 15 recommendations (**Table 4** and **Supplement 5**).

**Table 4.** Summary of the 15 strategic recommendations for developing and using an integrated national data source on potentially avoidable HIE-related brain injury. Supplement 5 details each recommendation.

| **Domain** | **Header of recommendation** |
| --- | --- |
| **Defining potentially avoidable brain injury to underpin an integrated data source** | 1. Use HIE as the core diagnostic descriptor while acknowledging the limitations of attributing causality<br>2. Focus on babies born at or beyond 36 weeks' gestation and ensure the definition captures mild cases of HIE |
| **Aligning data items across data sources and platforms** | 3. Consensus-build a catalogue for core data items for an integrated data source<br>4. Develop a shared data dictionary to support interoperability across existing data sources.<br>5. Standardise and embed clinical definitions and diagnostic thresholds for data items prone to various interpretations<br>6. Strengthen procurement standards for EPR systems to ensure interoperability and alignment with national datasets.<br>7. Configure local EPR systems to ensure consistency and interoperability |
| **Improving quality, reliability and feasibility of capturing data** | 8. Support contemporaneous, workflow-integrated data capture through co-designed systems<br>9. Explore the potential of technology-supported interpretation tools to improve consistency in data capture |
| **Enhancing linkage of data sources and long-term follow-up** | 10. Enable secure, scalable data linkage using a consistent encrypted pseudonymised identifier<br>11. Explore the feasibility of extending structured follow-up and data collection beyond two years for babies with suspected or confirmed brain injury. |
| **Strengthening data-driven feedback mechanisms** | 12. Align data feedback mechanisms with existing infrastructure to support local improvement<br>13. Consider dedicated support and capability-building for high-quality data capture and use |
| **Improving data and data sharing relevant to people accessing maternity or neonatal services** | 14. Explore the inclusion of new data items that reflect feelings of safety, continuity of intrapartum care, and responsiveness to concerns during perinatal care of any population<br>15. Co-design approaches for communicating brain injury-related data to share data with families in ways that support timely understanding, informed choice, and trust |

## Discussion

This study presents the first co-created strategy for supporting data-driven efforts to reduce avoidable HIE-related brain injury around the time of birth. A data dictionary review and work with 450 professional and patient/family stakeholders generated 15 recommendations to address previously identified challenges of UK maternity and neonatal data systems, including inconsistent definitions, limited linkage across datasets, duplication in data entry practices, incomplete coverage of key data items, and limitations in data meaningful to families.^16,20,21^ The recommendations provide an ambitious yet practical roadmap for developing and using an integrated national data source comprising routinely collected maternity and neonatal data.

A key recommendation is to build on existing national datasets and data flows to create an integrated data source, rather than establishing a new data registry. The recommendations propose phased rollout, starting with a core set of existing items that can be linked across current datasets, alongside a strategic approach to improving the consistency and quality of this set. This could run in parallel with identifying and incorporating new items highlighted by our consultations, including under-reported or missing intrapartum risk factors and service users’ experiences of care. Methods used to develop Core Outcome Sets for neonatal research^34,35^ offer useful principles for involving both professionals and maternity service users in decisions about selecting, amending, or adding items to a core set.

Other recommendations support the development of a standardised definition of potentially avoidable brain injury around the time of birth, which is essential for an integrated data source.^1,22,36^ Although neonatal encephalopathy (NE) may be a more aetiologically neutral term to describe disturbed neurologic function shortly after birth,^36^ the use of HIE aligns with current national guidelines for therapeutic hypothermia^37^ and is more consistently identifiable in structured fields of existing datasets. Our findings indicate the definition should be pragmatic and sensitive to emerging trial and observational evidence on clinical indicators of HIE,^1^ ^7^ while addressing data gaps around mild HIE and focusing on at/near-term infants, given existing neonatal reporting for preterm babies.^7^

Our consultations highlighted that effective maternity and neonatal data systems require attention not only to technical infrastructure supporting contemporaneous, workflow⍰integrated data capture,^23,38^ but also to socio-technical features and organisational data culture that shape how information is recorded, interpreted, and used for improvement. Improving data entry systems through collaboration with frontline professionals, clinicians with data responsibilities, human factors specialists, EPR vendors, and families will be essential to ensure that data collection better fits within routine workflows and minimises burden.^39^ Equally important is collaborative development of feedback mechanisms that build on existing audit platforms (e.g. National Maternity and Perinatal Audit, National Neonatal Audit Programme), ensuring dashboards and analytic tools support local governance, structured quality improvement, and multidisciplinary engagement.^20,22,30–33^ Sustaining high-quality data capture and use also demands investment in capability-building, including protected time, training, and clear communication that frames data as a tool for safer care rather than administrative compliance. Without this, even sophisticated technical systems are unlikely to realise their potential. It is likely that

Another gap is the lack of established systems and principles for sharing and visualising relevant data with service users. Although preliminary work exists,^40^ our workshops highlighted the need for nationally coordinated approaches. Engagement with EPR vendors will be important to all of this.

### Strengths and limitations

Our approach enabled the expertise of diverse stakeholders to be mobilised at scale to agree strategic recommendations in an area of pressing need.^22,23^ While focused on UK perinatal brain injury, the approach and findings may be highly relevant to address data challenges and improvement needs in other maternity and neonatal areas (both in the UK and beyond),^17,21,24–26^ though this requires further evaluation.

A limitation is that it was not feasible to involve all relevant stakeholders. Convenience sampling for the national survey supported broad inclusion but not full representativeness, while future efforts should engage additional senior UK decision makers beyond those consulted in the multidisciplinary groups. A further technical issue to address is understanding how any updates to, or stronger linkage within, existing datasets may affect the identification of missing items identified in our earlier data dictionary review.^22^ Finally, we acknowledge that many of the proposed recommendations are aspirational, but also believe that without a clear improvement vision it will be difficult to gain traction and make change over the long-term.

### Conclusions

The co-created recommendations offer an ambitious yet practical roadmap for developing an integrated UK data source focused on potentially avoidable HIE-related brain injury, building on existing national dataset infrastructure. Even a basic version of an integrated data source would already help inform efforts to support safer maternity care, enable continuous improvement cycles, and guide future investment in digital infrastructure.

## Acknowledgements

We deeply honour the contributions of Professor Tim Draycott (1964* – 2025**^†^**) to the presented work and his other invaluable contributions to maternity safety in the UK and beyond.

We are grateful for the many and varied contributions from colleagues across the Avoidable Brain Injury in Childbirth (ABC) programme team (https://www.rcog.org.uk/about-us/quality-improvement-clinical-audit-and-research-projects/avoiding-brain-injury-in-childbirth-abc/) and external to the team.

We thank the Thiscovery team (https://www.thiscovery.org/) for their range of contributions to the online infrastructure survey and forum-based consultations

Contributor Groups

- **ABC Data Advisory Group members:** Aarti Mistry, Alessandra Morelli, Alissa Fremeaux, Annette Anderson, Asma Khalil, Chris Gale, David Odd, Ela Chakkarapani, Elaine Boyle, George Dunn, Hilary Cruikshank, Humfrey Legge, Ian Gallimore, Jane Sandall, Jules Gudgeon, Julie-Clare Becher, Lara Shipley, Neil Muchatuta, Oliver Rackham, Philip Steer, Philippa Rees, Rachel Walsh, Sam Oddie, Sara Webb, Sarah Hookes, Sarah Seaton, Sian Harrington, Tim Draycott, Tim van Hasselt, Tng Chang Kwok
- **ABC PPI group**: Cate Maddison, Julia Clark, Heather Simmonds-Copete, Penny Harger, Sarah Fisher, Sarah Seddon

## Consent

Ethical review for the survey was sought through the University of Cambridge Psychology Research Ethics Committee (PRE.2024.087).

## Funding

The Avoiding Brain Injury in Childbirth programme (Department of Health and Social Care, UK) supported this work. THIS Institute is funded by the Health Foundation, Grant/Award Number: RHZF/001 - RG88620. The Health Foundation is an independent charity committed to bringing about better health and health care for people in the UK.

## Disclosure of interest

All authors and contributor group members declare they have no competing interests.

## Data availability statement

The data that support the findings of this evaluation are available on request from the corresponding author. Access to fully anonymized data may be granted to bona fide researchers under a data sharing agreement. The data are not publicly available because of privacy or ethical restrictions.

## Contribution to Authorship

Jan W. van der Scheer is the guarantor for this article. All authors read and approved the final manuscript. Their specific contributions, following CRediT (Contributor Roles Taxonomy), are as follows:

Jan W. van der Scheer contributed to the conceptualisation, funding acquisition, formal analysis, investigation, methodology, project administration, writing – original draft preparation, and writing – review and editing.

Kirstin Webster contributed to the investigation, methodology, project administration, writing – original draft preparation, and writing – review and editing.

Muhammed Ally Hussein Wahedally contributed to the formal analysis, investigation, and writing – review and editing.

Jane O’Hara contributed to the methodology, project administration, supervision, and writing – review and editing.

Tim Draycott contributed to the conceptualization, funding acquisition, methodology, project administration, supervision, and writing – review and editing

Andrew Demitri contributed to the investigation, project administration, and writing – review and editing.

Rachna Bahl contributed to the methodology, project administration, supervision, and writing – review and editing.

Mary Dixon-Woods contributed to the conceptualisation, funding acquisition, methodology, project administration, supervision, and writing – review and editing.

## Notes

### Competing Interest Statement

The authors have declared no competing interest.

### Author Declarations

Ethical approval for the survey was given by the University of Cambridge Psychology Research Ethics Committee (PRE.2024.087, see https://www.bio.cam.ac.uk/psyres for further information).

