## Supplement 1 for "Improving UK data on avoidable perinatal brain injury: consultations and co-creation of recommendations"

### Supplement 1. Survey methods

#### Overview of survey methods

We conducted an online survey using Thiscovery (<https://thiscovery.org/>), a secure platform designed to support collaborative work on health and care improvement. Eligible participants included UK-based clinicians working in maternity or neonatal care. Ethical review was sought through the University of Cambridge Psychology Research Ethics Committee (PRE.2024.087).

Participants were recruited via professional networks and social media, with informed consent obtained electronically prior to participation. Convenience sampling was used, with no minimum or maximum specified sample size, leading to a sample of **411 professionals from all UK regions**, including roughly 70% maternity (consultant and trainee obstetricians, senior and junior midwives, and midwives with digital roles) and 30% neonatal (consultant and trainee neonatologists, advanced neonatal nurse practitioners, and neonatal allied health professionals).

The survey gathered views on: (i) defining potentially avoidable brain injury, (ii) data items for a future integrated data source, and (iii) using routinely collected data to support improvements in care. Survey questions (see **below**) were developed iteratively with the authorial team and through pilot testing with four maternity and neonatal professionals.

We analysed closed-ended responses using descriptive statistics, including data from all participants who completed at least one closed-ended question. Free-text responses were categorised in a matrix alongside aggregated responses for each closed-ended question to identify patterns across topics and then thematically summarised (O’Cathain et al, 2010).

- O'Cathain A, Murphy E, Nicholl J. Three techniques for integrating data in mixed methods studies. Bmj. 2010 Sep 17;341:c4587.

#### Questions used in the survey

##### Definition

A clear and agreed clinical definition of avoidable brain injury is very important for a dataset dedicated to monitoring clinical indicators and risk of factors of avoidable brain injury. We’d like to ask your help with developing this.

**Which of the following best describes the clinical condition of avoidable brain injury around the time of birth?** Please select one or more that apply.

- Neonatal encephalopathy (NE)
- Hypoxic-ischaemic encephalopathy (HIE)
- A brain injury related to a series of fetal and neonatal insults around the time of birth
- A brain injury that happens around the time of birth when oxygen to the brain is reduced or stopped
- Other, please describe: [free-text]
- I don’t know [unique answer option]

**Which of the following best describes avoidability of brain injury around the time of birth?** Please select one or more that apply.

- Injury that has any avoidable cause
- Injury that may be avoidable under conditions of optimal perinatal care
- Injury that is caused by suboptimal perinatal care
- Other, please describe: [free-text]
- I don’t know [unique answer option]

**Which of the following best describes the consequences of brain injury around the time of birth?** Please select one or more that apply.

- Injury resulting in neurological impairments
- Injury resulting in developmental impairments
- Injury resulting in functional impairments
- Other, please describe: [free-text]
- I don’t know [unique answer option]

**Please share below any comments about a proposed clinical definition of avoidable brain injury around the time of birth.**

##### Data items

You’re probably asked to enter data into a local electronic patient record (EPR) as part of your clinical practice. Some – but not all – of these EPR data are later extracted into national digital datasets.

Examples of national digital datasets are the Maternity Services Data Set (MSDS), the Hospital Episode Statistics for Admitted Patient Care (HES APC), and the National Neonatal Research Database (NNRD). We’ve reviewed these national datasets and have identified data items that may be relevant to avoidable brain injury.

We’d like to ask your views on the importance of various types of data items, what’s missing in current datasets, and issues with accurately capturing data during clinical practice. Your views will inform a selection of the most important items, with the aim to bring these together in a consistent way in a single national database dedicated to avoidable brain injury.

###### Importance of data items

**In your view, how important are the following types of data items when trying to understand the *frequency* of avoidable brain injury around the time of birth?** There are 4 items on this page. You can skip an item if you don’t have a view or can’t give an answer.

(1) Clinical status at birth (e.g. Apgar score, need for resuscitation, umbilical cord bloods)

Not at all important (1) – (2) – (3) – (4) – (5) Extremely important

(2) Post-natal neurological assessment (e.g. central tone, consciousness, cerebral function monitoring [CFAM])

Not at all important (1) – (2) – (3) – (4) – (5) Extremely important

(3) Interventions that indicate efforts were made to avoid or reduce the impact of brain injury (e.g. transfer to neonatal unit, anticonvulsant therapy, passive or active therapeutic hypothermia [“cooling”])

Not at all important (1) – (2) – (3) – (4) – (5) Extremely important

(4) Brain imaging outcomes (e.g. MRI, EEG, cranial ultrasound)

Not at all important (1) – (2) – (3) – (4) – (5) Extremely important

**Please share below if you have any comments about the answers you provided above.**

**In your view, how important are the following types of data items when trying to understand *clinical risks* associated with avoidable brain injury around the time of birth?** There are 4 items on this page. You can skip an item if you don’t have a view or can’t give an answer.

(1) Maternal characteristics (e.g. ethnic group, socioeconomic deprivation, maternal age)

Not at all important (1) – (2) – (3) – (4) – (5) Extremely important

(2) Pre-existing risk factors (e.g. BMI at first booking, pre-existing hypertension, pre-existing diabetes, smoking)

Not at all important (1) – (2) – (3) – (4) – (5) Extremely important

(3) Antenatal risk factors (e.g. suspected fetal growth restriction, pre-eclampsia, gestational diabetes or hypertension)

Not at all important (1) – (2) – (3) – (4) – (5) Extremely important

(4) Intrapartum risk factors (e.g. gestational age, duration of labour, maternal pyrexia, meconium)

Not at all important (1) – (2) – (3) – (4) – (5) Extremely important

**Please share below if you have any comments about the answers you provided above.**

###### New data items for national datasets

Next, we’d like to ask your views on types of data related to avoidable brain injury that **aren’t** currently captured in existing national datasets.

Below are examples of data that many units record in their local electronic patient records (EPR), but that are currently not extracted into the national datasets.

**Which of the following data items do you think would be important to extract into existing national datasets?** Please select one or more options.

- Whether intrapartum intermittent auscultation (IA) or cardiotocography (CTG) was used
- How intrapartum CTG was classified
- Recording of fetal movements
- Use and timing of cord clamping
- Whether fetal scalp blood testing was undertaken
- Whether shoulder dystocia occurred
- Whether placental abruption occurred
- Sarnat score and/or Thompson Encephalopathy Score
- Whether persistent pulmonary hypertension of the newborn (PPHN) developed
- None of the above
- Other: [free text]

###### Issues in capturing data items

**In your view, how difficult is it to accurately capture the following data items?** By “capturing”, we mean collecting the data item and recording it in an electronic patient record (EPR) during or after care provision.

(1) Ethnic group

Not difficult at all (1) – (2) – (3) – (4) – (5) Extremely difficult

(2) Pre-eclampsia

Not difficult at all (1) – (2) – (3) – (4) – (5) Extremely difficult

(3) Duration of second stage labour

Not difficult at all (1) – (2) – (3) – (4) – (5) Extremely difficult

(4) Apgar score at five minutes

Not difficult at all (1) – (2) – (3) – (4) – (5) Extremely difficult

(5) Umbilical cord pH

Not difficult at all (1) – (2) – (3) – (4) – (5) Extremely difficult

**What might help in accurately capturing data related to avoidable brain injury around the time of birth?**

##### Sharing data insights

Capturing data relevant to avoidable brain injury around the time of birth is only meaningful if the data can be used to improve practice. We’d like to ask your views on how best to share data insights.

**When thinking about your clinical practice and experiences, to what extent do you agree with the following statements?**

(1) Maternity/neonatal professionals should get better insights into summarised data on avoidable brain injury in their own unit

Strongly disagree (1) – Disagree (2) – Neither agree nor disagree (3) – Agree (4) – Strongly agree (5)

(2) Maternity/neonatal professionals are more likely to improve their practice if data on avoidable brain injury in their own unit were presented in an accessible and meaningful way.

Strongly disagree (1) – Disagree (2) – Neither agree nor disagree (3) – Agree (4) – Strongly agree (5)

(3) Maternity/neonatal professionals need further training on how to improve their practice using data on avoidable brain injury of their own unit

Strongly disagree (1) – Disagree (2) – Neither agree nor disagree (3) – Agree (4) – Strongly agree (5)

(4) Parents and people who are planning to be parents should have access to summarised data on avoidable brain injury in the unit they’re attending

Strongly disagree (1) – Disagree (2) – Neither agree nor disagree (3) – Agree (4) – Strongly agree (5)

(5) It’s important for all units to routinely monitor and review their own data on avoidable brain injury

Strongly disagree (1) – Disagree (2) – Neither agree nor disagree (3) – Agree (4) – Strongly agree (5)

##### Final reflections

**Is there anything else you’d like to share about using data on avoidable brain injury to improve practice?**

##### About you

Thank you so much for all your answers so far. Below are a few last questions to help us understand your professional role. After that, you can choose to share your email address for further project updates.

If you would like to skip sharing anything about your professional role, please tick the box and then select "Next" at the bottom of the page.

- Skip this section

**What is your professional role?**

- Maternity professional
- Neonatal professional
- Other, please describe: [free-text box]
- Prefer not to say

[If maternity professional was selected, the following question appears:]

**Please select one of the following options best describing your role:**

- Midwife band 5
- Midwife band 6
- Midwife band 7
- Midwife band 8
- Student Midwife
- Maternity Support Worker / Healthcare Assistant
- Registered Nurse/Midwife not Qualified in Specialty (not-QIS)
- Registered Nurse/Midwife Qualified in Specialty (QIS)
- Physician Associate
- Speciality trainee ST1-2
- Speciality trainee ST3-5
- Speciality trainee ST6+
- SAS (Specialist, Associate Specialist and Specialty) doctor
- Consultant Obstetrician
- Other (please describe): [free text]
- Prefer not to say

[If neonatal professional was selected, the following question appears:]

**Please select one of the following options best describing your role:**

- Registered Nurse/Midwife not Qualified in Specialty (not-QIS)
- Registered Nurse/Midwife Qualified in Specialty (QIS)
- Nursery Nurse/Healthcare Assistant
- Transport Nurse
- Enhanced Neonatal Nurse Practitioner (ENNP)
- Advanced Neonatal Nurse Practitioner (ANNP)
- Neonatal Allied Health Professional (AHP)
- Physician Associate
- Paediatric specialty trainee ST1-3
- Paediatric specialty trainee ST4-5
- Neonatal GRID trainee ST6+
- SAS (Specialist, Associate Specialist and Specialty) doctor
- Consultant neonatologist
- Other (please describe): [free text]
- Prefer not to say

**How long have you worked in maternity or neonatal care?**

- 1-3 years
- 4-5 years
- 6-10 years
- 11-20 years
- More than 20 years
- Prefer not to say

**Do you hold a clinical leadership role with responsibilities for the quality of routinely collected data across your unit?**

- No
- Yes, please describe: [free text]
- Prefer not to say

**What type of maternity unit or setting do you currently work in?** If you work in more than one setting, please select all options that apply.

- Neonatal unit
- Obstetric unit
- Alongside midwifery unit (a midwifery-led unit or birth centre situated in the same hospital or on the same site as an obstetric unit)
- Freestanding midwifery unit (a midwifery-led unit or birth centre not situated in a hospital or site with an obstetric unit)
- Community
- Other (please describe): [free-text]
- Prefer not to say

**What region in the UK do you (mostly) work in?**

- East of England
- London
- Midlands
- North East and Yorkshire
- North West
- Northern Ireland
- Scotland
- South East
- South West
- Wales
- Prefer not to say
