## Supplement 2 for "Improving UK data on avoidable perinatal brain injury: consultations and co-creation of recommendations"

### Supplement 2. Multidisciplinary consultation: methods and findings

#### Overview of online discussion methods

We hosted a forum-based consultation on the Thiscovery platform (<https://thiscovery.org/>) – a secure platform designed to support collaborative work on health and care improvement – with a multidisciplinary advisory group. Members were purposively invited to ensure broad representation of the expertise required to address the complex clinical, technical, and policy dimensions of avoidable brain injury data in the UK. Invitations were based on contributions to earlier phases of the programme (van der Scheer et al, 2025) or recognition of domain-specific expertise. All were invited to be named in the authorial contributor group for the outputs of the study.

The advisory group included **35 individuals with leadership roles or expertise** spanning obstetrics, midwifery, neonatology, paediatrics, anaesthetics, clinical audit, epidemiology, statistics, safety investigations, and medical litigation.

The consultation was organised around the following topics: (i) inform defining avoidable brain injury in the perinatal context, (ii) assess and recommend data items for an integrated source, and (ii) review draft recommendations (**see below**) that drew on issues identified in the data dictionary review and interview consultation (van der Scheer et al, 2025). Each topic was introduced using background briefing documents that incorporated findings from the national survey, summarised key literature, and outlined discussion questions. Contributions were submitted via short surveys, together with the option for further exploration through asynchronous forum discussions among group members.

Responses were summarised using a deductive approach informed by the Framework Method (Gale et al, 2013) and structured around predefined topic areas: definitions, data items, standardisation, linkage, quality, feedback to professionals, and feedback to families (van der Scheer et al, 2025).

- Gale NK, Heath G, Cameron E, Rashid S, Redwood S. Using the framework method for the analysis of qualitative data in multi-disciplinary health research. BMC Medical Research Methodology. 2013;13(1):117.
- van der Scheer JW, et al. Improving UK data on avoidable perinatal brain injury: review of data dictionaries and consultation. Pediatr Res. 2025 Jan 30.

#### Set of 15 draft recommendations that the multidisciplinary advisory group was asked to rate and review

1. **Coordinate national oversight through a shared data dictionary** – National bodies responsible for maternity and neonatal data should jointly oversee the alignment of brain injury-relevant data by developing and maintaining a shared data dictionary for an agreed catalogue of items. This should include agreement on standardised data definitions, consistent use of classification systems—such as the Systematized Nomenclature of Medicine Clinical Terms (SNOMED CT), the International Statistical Classification of Diseases and Related Health Problems 10^th^ Revision (ICD-10) and the Office of Population Censuses and Surveys' Classification of Surgical Operations version 4 (OPCS-4)—and development of metadata conventions such as naming, formats, and time-stamping rules to ensure interoperability across maternity and neonatal datasets. The governance structure should define clear responsibilities for maintaining and updating definitions and dataset specifications, and ensure that future versions of national datasets are explicitly designed to support reliable linkage and meaningful comparison of brain-injury relevant data.
2. **Standardise and embed clinical definitions for high-variability data items** – To improve the reliability of brain injury-relevant data, clinical definitions and classification criteria for commonly variable data items—such as fetal heart rate features and eligibility for therapeutic hypothermia—should be clearly defined, nationally standardised, and embedded into guidance and training. The focus should be on clinical accuracy and usability, with definitions developed collaboratively across maternity and neonatal disciplines and embedded in training, clinical tools, and data specifications. Where appropriate, the specification of definitions and criteria should support contemporaneous data capture during routine care.
3. **Ensure all EPR platforms support structured, standards-aligned capture of brain injury data** – To minimise inconsistency, electronic patient record (EPR) platforms should support the capture of brain injury-relevant data items using structured fields aligned with nationally agreed clinical definitions, classification systems (e.g. SNOMED CT, ICD-10, OPCS-4), and metadata conventions (e.g. naming, formats, time-stamping). Platforms should minimise reliance on free-text input and enable data entry that is interoperable and suitable for linkage, audit, and analysis.
4. **Set clear procurement expectations to ensure interoperability and data standards** – To reduce data inconsistencies and enable linkage across systems, procurement processes must require that EPR platforms support structured capture of brain injury-relevant data and export data in interoperable formats (i.e. formats that allow systems to exchange and interpret data consistently). Specifications should mandate alignment with nationally agreed definitions and classification systems (e.g. SNOMED CT, ICD-10, OPCS-4) and ensure compliance with national datasets (e.g. MSDS, HES APC, Neonatal Data Set [NDS]).
5. **Configure local systems to support semantic interoperability at the point of care** – To support consistency at the point of care, local EPR configurations should enable structured, clinically relevant capture of brain injury data using nationally agreed clinical definitions and classification systems (e.g. SNOMED CT, ICD-10, OPCS-4), while ensuring compliance with current national datasets (e.g. MSDS, HES APC and NDS). Local build decisions must prioritise the minimisation of free-text and maximise the use of codified data structures that enable semantic interoperability (i.e. the ability of systems to not only exchange data but understand it in the same way).
6. **Prioritise contemporaneous data capture through co-designed systems**– To improve data quality and reduce professional burden, data should be recorded contemporaneously—in real time as part of routine care—rather than retrospectively. The design of systems that support contemporaneous data collection should involve end-users (e.g. frontline staff), data managers (e.g. digital midwives), and other specialists (e.g. human factors engineers, EPR specialists). Systems should draw on successful data flow models from other NHS settings and avoid reliance on handwritten or free-text entries where structured options are viable.
7. **Design intuitive, workflow-integrated data entry interfaces** – To help reduce variation in how clinicians record key data items, electronic data entry systems should incorporate structured fields with decision-support features—such as dropdown menus, pre-populated values, auto-calculations, and prompts for action that are triggered only when clinically relevant. Interface design should prioritise usability and integration into clinical workflows to ensure that data capture happens as part of routine care rather than as an add-on.
8. **Use technology and structured tools to reduce subjectivity and improve interpretive consistency** – To further enhance intra- and inter-rater reliability, national bodies responsible for maternity and neonatal data should consider technology-driven solutions to support the consistent interpretation of complex clinical information—such as automated analysis of fetal monitoring, standardised EEG/MRI reporting, and natural language processing to structure narrative data where it must be used. Where full automation is not yet feasible or reliable, the use of structured interpretation tools, validated templates, and standardised classification systems may support clinical judgement, reduce variability across clinicians and trusts, and facilitate high-quality, comparable data capture at scale.
9. **Enable secure, scalable data linkage using a consistent encrypted pseudonymised identifier** – An effective, efficient and privacy-preserving method for linking relevant data items across maternity and neonatal datasets—such as the Maternity Services Data Set (MSDS), Hospital Episode Statistics for Admitted Patient Care (HES APC), and the Neonatal Data Set (NDS)—is to use a consistent, encrypted pseudonymised identifier. This approach enables person-level linkage of data captured through national audit programmes such as the National Maternity and Perinatal Audit (NMPA) and the National Neonatal Audit Programme (NNAP), while protecting individual privacy and supporting integration across data sources.
10. **Extend follow-up data collection beyond two years for babies and children with suspected or confirmed brain injury** – To understand the long-term impact of perinatal brain injury, data collection should extend beyond the standard two-year neonatal follow-up. This includes capturing structured data from each neonatal follow-up clinic appointment, community health services (e.g. health visitors), and paediatric services, including referrals, assessments, and interventions. Expanding the time horizon of data collection will support more complete identification of developmental challenges, service needs of the child, and outcomes that may only become apparent later in childhood.
11. **Develop longitudinal data infrastructure to link maternity, neonatal, and paediatric records –** To support long-term follow-up of babies and children with suspected or confirmed brain injury, national infrastructure should be developed to enable secure linkage between perinatal, paediatric and community datasets. This includes connecting datasets such as the Maternity Services Data Set (MSDS), Neonatal Data Set (NDS), Community Services Data Set (CSDS), Child Health Information Services (CHIS), Paediatric Critical Care Minimum Data Set (PCCMDS)

and, where appropriate, education or disability support systems. Implementation will require standardised identifiers across systems, robust data sharing agreements, and interoperable platforms capable of linking data across organisational boundaries. Creating this longitudinal view will support early intervention, care coordination, and long-term evaluation of outcomes.

12. **Strengthen digital feedback systems through secure, role-specific dashboards with timely data access** – To support timely action to reduce risk of perinatal brain injury, national audit programmes such as the National Maternity and Perinatal Audit (NMPA) and National Neonatal Audit Programme (NNAP) should be enabled to evolve their existing reporting into interactive dashboards that provide secure, role-specific and timely access to brain injury-related data at trust level. While public dashboards must display aggregated data with a delay, secure-access dashboards could, over time and where data flows allow, offer near real-time insights without compromising patient confidentiality. This could begin by incorporating key brain injury-relevant data into existing dashboards, with authorised log-in functionality for trusts. Role-specific views should allow clinicians to review patient-level lists, clinical leads to track unit-level trends, and managers to monitor regional and national benchmarking. This infrastructure can then be expanded gradually to support more advanced visualisations and integration with local, regional, and national quality improvement cycles.
13. **Embed simple, actionable prompts within feedback platforms** – Embedding clear, feasible action planning tools within feedback systems increases the likelihood that teams respond effectively to the data, even in busy and resource-constrained environments. Audit and feedback infrastructure should go beyond passive reporting by incorporating simple, context-specific action prompts relevant to reducing risk of avoidable brain injury. This might include automated reminders for reviewing cases, prompts for improving documentation of risk factors or clinical indicators of brain injury, or direct links to local protocols and national guidelines. These tools should be co-designed and piloted with maternity and neonatal teams to ensure they support—not overwhelm—clinical workflows.
14. **Integrate data-driven feedback mechanisms into routine governance and quality improvement structures** – Efforts to reduce risk of avoidable brain injury should build on existing audit infrastructure by ensuring that relevant indicators are routinely reviewed within multidisciplinary governance processes. NMPA and NNAP are well positioned to support integration of feedback into local improvement cycles, enabling structured reviews of trends, causes, and system-level factors contributing to brain injury. Making this actionable would require data-driven feedback to be integrated into local governance forums such as multidisciplinary meetings and perinatal morbidity reviews, with clear roles and responsibilities for acting on insights. Ongoing co-design with clinical teams will be key to ensuring feedback remains meaningful, credible, and manageable. In time, feedback systems could be embedded into clinical systems for real-time use, helping data-driven feedback become part of everyday practice rather than an external process.
15. **Consider dedicated support and capability-building for high-quality data capture and use** – To embed data-driven improvement into routine practice, national workforce strategies should consider how maternity and neonatal units could be supported with the protected time, training, and staffing to enable high-quality data capture and interpretation. This may include dedicated support for digital midwives, neonatal data leads, and other team members involved in using data to drive improvement—alongside opportunities to build capability of all relevant professionals in recording accurate data and interpreting local insights. Without the resources for professionals to engage meaningfully with data, even the most advanced systems will fall short of their potential to reduce avoidable harm.

#### Rating of the recommendations by 20 out of 35 members

| **Recommendation** | **n (Agree/Strongly agree)** | **% of 20** |
| --- | --- | --- |
| Recommendation 1 | 20 | 100.0% |
| Recommendation 2 | 19 | 95.0% |
| Recommendation 3 | 18 | 90.0% |
| Recommendation 4 | 17 | 85.0% |
| Recommendation 5 | 18 | 90.0% |
| Recommendation 6 | 20 | 100.0% |
| Recommendation 7 | 20 | 100.0% |
| Recommendation 8 | 15 | 75.0% |
| Recommendation 9 | 16 | 80.0% |
| Recommendation 10 | 17 | 85.0% |
| Recommendation 11 | 16 | 80.0% |
| Recommendation 12 | 16 | 80.0% |
| Recommendation 13 | 17 | 85.0% |
| Recommendation 14 | 20 | 100.0% |
| Recommendation 15 | 17 | 85.0% |

#### Summary of feedback on draft recommendations

Key free-text feedback on the draft recommendations included:

- Add that a national regulatory mechanism is needed to formally require vendors and providers to adhere to agreed national definitions, classifications, and metadata standards so that consistent and standardised data collection is assured across systems.
- Ensure alignment with other current or upcoming national programmes such as NHS England’s Maternity Outcome Support System (MOSS) and NHS Resolution’s Early Notification Scheme
- Emphasise that system design should incorporate meaningful involvement of clinicians, patients, and parents, including options for incorporating patient‑entered data where appropriate.
- Highlight that data infrastructure should enable integration between EPRs and clinical devices to streamline real‑time data capture and reduce manual transcription.
- Emphasise that secure but practical mechanisms are needed to support pseudonymised data linkage while maintaining robust safeguards, transparent access pathways, and appropriate consent for secondary uses.
- Clarify that extending follow‑up requires addressing fundamental variability in follow‑up practices across services, as well as challenges in linking datasets consistently over time.
- Highlight that public‑facing dashboards require careful consideration to avoid misinterpretation, unintended behavioural effects, or risks of identifiability where case numbers are small.
- Show that improvements in data capture and use depend on broader system conditions, including adequate staffing, digital expertise, protected time, and embedding responsibilities within existing roles.
- Recommend that implementing an updated national data dictionary would benefit from phased, coordinated development that supports alignment with existing data initiatives and reduces supplier‑related delays.
- Emphasise that effective uptake of improved data systems relies on intuitive design, workflow integration, and sufficient training so that available structured features are reliably used in practice.
