## Supplement 3 for "Improving UK data on avoidable perinatal brain injury: consultations and co-creation of recommendations"

### Supplement 3. Suggested data items in survey, multidisciplinary consultation and patient/family workshops

#### Data items identified as important that may not currently be captured in national datasets: pre-selected responses of survey participants

Survey participants’ responses (*N*=367) to selecting a range of pre-specified data items – identified in our previous data dictionary review as items that may not be captured in existing national datasets (van der Scheer et al, 2025) – as important for an integrated data source.

| **Example data item** | ***n*** | **%** |
| --- | --- | --- |
| Whether placental abruption occurred | 329 | 89.6% |
| Whether shoulder dystocia occurred | 327 | 89.1% |
| How intrapartum CTG was classified | 309 | 84.2% |
| Whether intrapartum IA or cardiotocography CTG was used | 293 | 79.8% |
| Recording of fetal movements | 227 | 61.9% |
| Use and timing of cord clamping | 176 | 48.0% |
| Sarnat score and/or Thompson Encephalopathy Score | 144 | 39.2% |
| Whether fetal scalp blood testing was undertaken | 135 | 36.8% |
| Whether persistent pulmonary hypertension of the newborn developed | 126 | 34.3% |
| Other, please describe | 34 | 9.3% |
| None of the above | 2 | 0.5% |

CTG = cardiotocography; IA = intermittent auscultation

#### Data items identified as important that may not currently be captured in national datasets: free-text suggestions from survey participants and advisory group members.

| **Category** | **Suggested additional data items** |
| --- | --- |
| **Clinical indicators** |  |
| Clinical status at or shortly after birth | - Neurological assessment against cooling criteria - Multi-organ failure - Dysglycaemia |
| Interventions that indicate efforts were made to avoid or reduce the impact of brain injury | - Involvement of allied health professionals during neonatal intensive care - Type of cooling available at the birth centre and via transport (passive or active) - Referral time to neonatal retrieval services and time to arrival of the transport team - Mode of transport and journey duration for uplift transfers (road or air) - Time to initiation of therapeutic hypothermia - Age at which therapeutic temperature was achieved - Therapeutic temperature maintenance during transfer, recorded at departure and arrival at tertiary cooling centre - Instances of therapeutic hypothermia discontinued prior to 72 hours - Length of neonatal intensive care unit (NICU) stay |
| Post-natal assessments | - Neonatal withdrawal scores (e.g. Finnegan Score) - Cranial ultrasound of preterm infants - Strokes, bleeds and trauma imaged with computed tomography or ultrasound imaging - Results from placental histology |
| Developmental follow-up assessments | - Prechtl General Movement assessment - Hammersmith assessment - Neurodevelopmental outcome at 2 years of age |
| **Risk factors** |  |
| Maternal characteristics or risk factors | - Alcohol consumption |
| Antenatal risk factors | - Multiple pregnancy - Presence of severe maternal morbidity during the current pregnancy - Estimated fetal weight or growth centile (including large for gestational age) - Concerns about antenatal fetal movements - Care outside clinical guidance |
| Intrapartum risk factors | - Birth outside national guidance (e.g. birth location, monitoring practices) - Induction or augmentation of labour (including method used and use of oxytocin) - Reason for induction or augmentation - Concerns about intrapartum fetal movements - Duration of fetal heart rate abnormality - CTG classification accuracy - Actions following escalation based on CTG classification - Fetal blood sampling and base deficit values - Treatment for uterine hyperstimulation - Mode of birth - Urgency of delivery (e.g. caesarean birth category) - Decision-to-delivery interval (instrumental or caesarean) - Use and complications of anaesthesia during delivery - Presence of impacted fetal head during caesarean birth - Uterine rupture - Amniotic fluid embolism - Cord prolapse - Fetal haemorrhage |

#### Data items identified as important that may not currently be captured in national datasets: suggestions from patient/ family representatives

Suggested types of data items by patient and public representatives (n=9) to capture in national data sources

| **Type of data item** | **Example of suggested data items** | **Illustrative quote** |
| --- | --- | --- |
| One-to-one care | Whether those in labour received continuous one-to-one care | *“Identifying gaps in one to one care would allow correlation of staffing levels/models to avoidable harm”* (Maternity safety advocate) |
| Feelings of safety | Whether those in labour felt physically and emotionally safe | *“What is important here is whether people feel safe - not just whether they are in fact safe.”* (Maternity service user) |
| Acknowledgement of concerns | Whether concerns raised by expectant parents were acknowledged and acted on by professionals during antenatal care or intrapartum care | *“[I]t wouldn’t be a problem if someone didn’t have any concerns so didn’t raise any, however if they did have concerns but didn’t feel able to raise them then this is an issue”* (Maternity service user) |
| Antenatal diagnostic opportunities | Whether antenatal diagnostic opportunities (e.g. screening, investigations, or referrals) were missed, delayed, or not acted upon | *“Antenatally, missed diagnostic windows - the nuance that clinical notes may lack.”* (Maternity safety advocate) |

van der Scheer JW, et al. Improving UK data on avoidable perinatal brain injury: review of data dictionaries and consultation. Pediatr Res. 2025 Jan 30.
