## Supplement 4 for "Improving UK data on avoidable perinatal brain injury: consultations and co-creation of recommendations"

### Supplement 4. Workshops with patient and public representatives

#### Methods

We facilitated three online workshops to engage people who had experience of UK maternity services as well as advocacy group representatives from charities supporting women and families affected by perinatal injury. **Nine women with experience of UK maternity services took part across the three workshops, including maternity safety advocates and representatives from charities supporting women and families** (particularly those affected by perinatal injury and those whose voices are under-represented). All were part of established patient and public involvement or advocacy groups, and were invited to be named in the authorial contributor group for the outputs of the study.

Facilitated by a healthcare improvement researcher (JWvdS) and a neonatal professional (KW), the semi-structured workshop discussions explored: (i) what types of brain injury-related data are most meaningful to families, (ii) how data should be shared to support decision-making, and (iii) what good communication looks like in the context of sensitive information.

Across the three workshops, discussions centred on real-life experiences, with the aim of identifying limitations in current data and generating ideas on information and feedback tools that could better support families during pregnancy, labour, and neonatal care. Preliminary findings from the national survey were shared during the sessions to prompt reflection, but otherwise the discussions were designed to be independent of the professional consultations. We summarised the facilitator notes from the workshops to identify recurring priorities and concerns related to family-facing aspects of data items and use.

#### Summary of findings in workshops

The following data summary with key findings was drafted based on the three workshops with patient and public representatives (n=9) and subsequently shared with them to receive further feedback and help shape recommendations with family-facing components:

- Some aspects of care that matter deeply to those who have given birth are not currently recorded in national datasets. These include whether those in labour received continuous one-to-one care, whether those in labour felt physically and emotionally safe, whether concerns raised by expectant parents were acknowledged and acted on by professionals during antenatal care or intrapartum care, and whether antenatal diagnostic opportunities (e.g. screening, investigations, or referrals) were missed, delayed, or not acted upon.
- Developing and testing structured data items in these areas could help identify patterns across services, strengthen trust between people accessing services and engaging with care providers, and ensure data better reflects the factors that contribute to feeling safe, heard, and supported. Such additions should be introduced thoughtfully, with attention to how they are asked and recorded, to ensure they are meaningful rather than tokenistic.
- How and when information is shared with families – especially during pregnancy – really matters. Contributors emphasised that communication about brain injury-related data must be handled with care, particularly at a time when people may already feel anxious or overwhelmed. Tools such as antenatal discussion prompts or better signposting to trusted information were welcomed as ways to support more informed and personalised conversations between people engaging with services and care providers
- Data must be accompanied by context in order to be meaningful. Numbers alone were seen as potentially confusing or even distressing – especially in services where people may have limited choices, such as where to give birth. People engaging with services likely want to understand not just “how many,” but also what was being done (or recommended to be done) to monitor and reduce safety risks in any given unit and incidence of specific conditions, and what support was available during and after care.
- The contributors highlighted the need for sharing contextualised data to inform people’s choices around available options such as those for screening, antenatal care, and birth plans. It was felt that those using maternity services also require insight into what support or care pathways are available prior to, during, and after birth to mitigate risk.
- For those who have experienced trauma or bereavement, priorities may differ. Contributors said those who have experienced trauma or bereavement would be more focused in knowing what has changed in response to past events (for example, how risks have now been mitigated), rather than seeing statistical data only.
- Capturing data not just on outcomes but on service-level factors – such as missed diagnoses, delays in response, or absent intrapartum monitoring – that help inform understanding of causation and prevention.
- Ensuring data shared with parents and expectant parents is timely, so that it supports informed choice and active participation in care decisions, rather than retrospective concern.
- Supporting people engaging with services in interpreting data, including access to contextualised information to move beyond raw numbers towards accessible and useful insights, such as explaining common contributory themes, clarifying action taken by services to address identified safety concerns in given units, personalised highlighting and explanation of data relevant to people’s specific circumstances and choices.
- Considering how information about prognosis and outcomes is made accessible to parents and expectant parents, to support them in planning care, advocating for services, and participating fully in decision-making.
- Overall, contributors emphasised that any approach to sharing data should reflect a range of lived experiences, and prioritise empathy, transparency, and trust-building.
