## Supplement 5 for "Improving UK data on avoidable perinatal brain injury: consultations and co-creation of recommendations"

### Supplement 5: Full outline of recommendations

#### Overview of methods to shape and agree recommendations

Following addressing the feedback on the draft recommendations (**Supplement 2**), the advisory group on a set of 13 recommendations around definitions, aligning data, linkage of data sources, data quality, feasibility of data capture, and data-driven feedback mechanisms for professionals (**recommendations 1 to 13 below**).

We summarised insights from the patient/family workshops (**Supplement 4**) and shared these with workshop attendees, together with proposed draft recommendations on capturing and sharing data with service users. Following iterative rounds of email-based revisions, the representatives agreed on two recommendations with family-facing components (**recommendation 14 and 15 below**).

#### Defining potentially avoidable brain injury to underpin an integrated data source

**1. Use HIE as the core diagnostic descriptor while acknowledging the limitations of attributing causality.**

To underpin a future integrated data source, national bodies should adopt a clinically meaningful, standardised definition of potentially avoidable brain injury around the time of birth, using hypoxic-ischaemic encephalopathy (HIE) as the core diagnostic descriptor. Although neonatal encephalopathy (NE) may be a more aetiologically neutral term to describe disturbed neurologic function shortly after birth, the use of HIE reflects current national guidelines (such as for therapeutic hypothermia) and is more consistently identifiable in structured fields within existing data sources. This approach should recognise the inherent challenges in determining precise causality in the early neonatal period.

**2. Focus on babies born at or beyond 36 weeks’ gestation and ensure the definition captures mild cases of HIE.**

Focusing the integrated data source on infants born at or beyond 36 weeks improves feasibility of accurate data capture by avoiding the greater complexity of brain injury in preterm babies, where causal pathways are even more multifactorial and often non-hypoxic. Moreover, preterm cases are already consistently captured in existing neonatal data reporting. Aligning the gestational threshold with the current national eligibility criteria for therapeutic hypothermia (≥36 weeks) also supports consistent case identification.

While national eligibility criteria for therapeutic hypothermia provide a pragmatic anchor for classifying moderate and severe cases, a definition of avoidable brain injury for the purposes of an integrated data source should also encompass babies with mild HIE. A significant number of these infants may experience long-term neurodevelopmental difficulties, but may not receive formal diagnoses, specialist review, or neonatal unit admission. This makes babies with mild HIE less likely to be systematically recorded in routine data, even though this is essential for understanding and reducing avoidable injury.

#### Aligning data items and digital systems to enable integrated data use

**3. Consensus-build a catalogue for core data items for an integrated data source.**

The development of an integrated data source should begin with a core set of data items already present in existing national datasets. This core set should be collaboratively agreed with key stakeholders, including professionals across maternity and neonatal services and patient and public representatives.

A practical starting point would be to generate a longlist of potential data items informed by previous consultations. This longlist can then be refined through a structured consensus process – drawing on Core Outcome Set methodology – to establish a final catalogue. Using Core Outcome Set methodology provides an established, patient-focused framework for ensuring that the views of women and families shape decisions about which items require improvement or inclusion.

Work to agree the core catalogue should run in parallel with efforts to identify and incorporate new data items. This includes addressing intrapartum risk factors and experiences of care, both of which are currently under-represented or absent in national datasets but are likely to be crucial for understanding potentially avoidable brain injury.

**4. Develop a shared data dictionary to support interoperability across existing data sources.**

To support interoperability across existing data sources, national bodies responsible for maternity and neonatal data should jointly oversee the development and maintenance of a shared data dictionary for an agreed catalogue of items relevant to brain injury. This should include clear data descriptions, guidance on use of coding systems (such as, where relevant, SNOMED CT, ICD-10, and OPCS-4), and standardised metadata conventions (such as naming, formats, and time-stamping).

A governance structure should define responsibilities for maintaining and updating data specifications, including input from clinicians and other users, with a clear mechanism for a phased rollout that starts with a core set of items. Any updates to data specifications should consider compatibility with historical data, so that longitudinal evaluations remain possible. This work could inform, and be aligned with, national data initiatives such as the Maternity Outcomes Signal System (MOSS).

**5. Standardise and embed clinical definitions and diagnostic thresholds for data items prone to various interpretations.**

To improve the reliability of brain injury-relevant data, clinical definitions and diagnostic thresholds should be clarified and standardised for data items that currently show significant variation in interpretation and recording, but are amenable to more consistent specification (such as the diagnosis of pre-eclampsia or eligibility for therapeutic hypothermia).

Definitions and thresholds should prioritise clinical accuracy and usability, be developed collaboratively across maternity and neonatal disciplines, and be embedded into data specifications. These should be supported, where appropriate, by structured interpretation tools, validated classification systems, and standardised reporting templates. For more complex items that are less amenable to full standardisation, such as fetal heart rate features, alternative strategies to support consistency in recording may be required (see also recommendation 10).

**6. Strengthen procurement standards for EPR systems to ensure interoperability and alignment with national datasets.**

Procurement processes for electronic patient record (EPR) systems should explicitly require structured, standards-aligned data capture to support the linkage and analysis of brain injury-relevant data. Contracts should mandate alignment with a nationally agreed data dictionary for brain injury and compatibility with national datasets. They should also require vendors to build in options to minimise reliance on free-text inputs and avoid duplicate data entries (allowing clinicians to record information once for multiple uses, such as documentation, audit, and analysis).

These expectations should be reinforced through national regulatory levers (e.g. Information Standards Notices) and supported by national procurement frameworks that empower trusts to select systems that meet functional requirements for integrated maternity and neonatal data use and minimise technical barriers to improvement.

**7. Configure local EPR systems to ensure consistency and interoperability**

Once procured, EPR systems must be locally configured to ensure alignment with a nationally agreed data dictionary for brain injury and datasets, minimise free-text inputs, and enable single data entries. Trusts should be supported with technical guidance to avoid local variation that undermines data quality or semantic interoperability (data retaining its meaning when exchanged between systems).

To help reduce variation in how clinicians record key data items, local EPR builds should incorporate structured fields with decision-support features, such as dropdown menus, pre-populated values, auto-calculations, and prompts for action that are triggered only when clinically relevant.

#### Improving quality, reliability and feasibility of capturing data

**8. Support contemporaneous, workflow-integrated data capture through co-designed systems.**

To improve data quality and reduce professional burden, data should be recorded contemporaneously – in real time as part of routine care – rather than retrospectively. The co-design of systems that support contemporaneous data collection should involve consultation and/or pilot testing with frontline staff, data managers such as digital midwives, human factors engineers, EPR specialists, and women and their birth partners.

These systems should systematically incorporate intuitive data entry interfaces (such as those already available in EPR systems), integrate bidirectional information streams where possible (such as between EPRs and medical equipment), and consider technological advances beyond current EPR capabilities (such as ambient voice technology). They should also draw on successful data flow models from other NHS settings and align with national data initiatives such as the Maternity Outcomes Signal System (MOSS).

**10. Explore the potential of technology-supported interpretation tools to improve consistency in data capture.**

National bodies, research funders, and EPR system vendors should explore and evaluate the use of emerging technologies – such as automated fetal monitoring interpretation, automated EEG reporting, and natural language processing – to reduce variation in how complex clinical data are interpreted and recorded. Although not yet widely feasible or validated for routine use, investment in this area could support longer-term improvements in data quality, and early pilot work or standardised templates could be developed to inform future scaling.

#### Enhancing linkage of data sources and long-term follow-up

**11. Enable secure, scalable data linkage using a consistent encrypted pseudonymised identifier.**

To support integrated analysis, reduce duplicative data collection, and enable long-term follow-up, a consistent encrypted pseudonymised identifier should be used across maternity and neonatal datasets – and, as a longer-term goal, across paediatric, community and related datasets (e.g. microbiology). This would allow privacy-preserving, person-level linkage of data from maternity and neonatal datasets through audit programmes such as the National Maternity and Perinatal Audit (NMPA) and National Neonatal Audit Programme (NNAP).

Secure mechanisms for access to linked data should be developed – such as through Accredited Researcher Status and Trusted Research Environments – with clear governance that maintains public trust, safeguards privacy, and enables accessibility to eligible analysts. Maximising the impact of data linkage will require improved digital maturity of EPR systems and standardisations described across recommendations 4 to 7.

**12. Explore the feasibility of extending structured follow-up and data collection beyond two years for babies with suspected or confirmed brain injury.**

Understanding the long-term impact of perinatal brain injury would benefit from extending structured follow-up and data collection within and beyond the current two-year window. Current follow-up practices vary widely across brain injury types, healthcare settings, and regions. As a first step, national bodies should explore the feasibility of developing more consistent pathways for follow-up and assess how data from later childhood – such as from community health services, paediatric clinics, or educational support – could be incorporated into linked maternity and neonatal data sources. In addition, parent-entered data collection could play an important role in complementing follow-up assessments.

#### Strengthening data-driven feedback mechanisms

**13. Align data feedback mechanisms with existing infrastructure to support local improvement.**

Dashboards and feedback mechanisms designed to help mitigate the risk of avoidable brain injury should build on existing audit infrastructure, particularly the National Maternity and Perinatal Audit (NMPA), the National Neonatal Audit Programme (NNAP), the MBRRACE-UK Real-Time Data Monitoring Tool, and the evolving Maternity Outcomes Signal System (MOSS). These systems should be designed to support local governance, perinatal morbidity reviews, and structured quality improvement processes, ensuring engagement with all relevant professionals.

Priority should be given to improving data quality and flow so that existing platforms can enable structured review of trends, contributory factors, and system-level issues at local, regional, and national levels – aligned with the needs of clinical, managerial, and other quality improvement audiences. Future enhancements may include action planning prompts or intelligent automation, but these must be co-designed with users and tested to ensure they are relevant, clear, and safe. Given the small number of cases, risks to anonymity, and the complexity of interpreting brain injury data, any public-facing dashboards should remain aggregated and presented with caution.

**14. Consider dedicated support and capability-building for high-quality data capture and use.**

Embedding data-driven improvement into routine practice will require national workforce strategies to address how maternity and neonatal services can be supported with protected time, training, and staffing to enable high-quality data capture and meaningful use. Dedicated training and clear communication of benefits (including positioning data as a tool for safer, better care rather than an administrative burden) should be co-designed with staff and endorsed by relevant Royal Colleges to support organisational and clinical buy-in.

These efforts should be complemented by additional support for digital midwives, neonatal data leads, and others who play a critical role in managing data for improvement. The focus of such additional support should be on empowering all members of the multidisciplinary team to confidently capture, interpret, and act on data. Without adequate time, skills, and support for professionals, even the most advanced data systems are unlikely to achieve their potential to reduce avoidable harm.

#### Improving data and data sharing relevant to people accessing maternity or neonatal services

**15. Explore the inclusion of new data items that reflect feelings of safety, continuity of intrapartum care, and responsiveness to concerns during perinatal care of any population**

National bodies responsible for maternity and neonatal datasets should consider including structured data items that capture and reflect important aspects of care currently under-represented in national datasets, including the needs, experiences and voices of families. This could include whether those in labour received continuous one-to-one care, whether they felt physically and emotionally safe, whether concerns were raised, acknowledged and acted on during antenatal or intrapartum care, whether opportunities for antenatal diagnosis were missed, and other data items. Including these types of measures may support families in antenatal decision-making about place of birth, provide deeper insight into care experiences, and help identify service-level contributory factors to brain injury.

Any new data items should be developed and tested in partnership with healthcare professionals, advocacy groups, and service users from diverse backgrounds to ensure they are insightful, feasible to capture, and supportive of care improvement. Particular attention should be paid to how and when questions are asked to service users, who asks those questions, and how responses are recorded and utilised, so that data gathering supports open, respectful, trust-building communication rather than causing distress, anxiety or harm.

Specific consideration should also be given to EDI (equality, diversity and inclusion) strategies in ensuring data is obtained from all groups and taking action to address any barriers to engagement in the provision of feedback about services.

**16. Co-design approaches for communicating brain injury-related data to help share data with families in ways that support timely understanding, informed choice, and trust**

National bodies responsible for maternity and neonatal services should engage with people accessing maternity and neonatal services, healthcare professionals, and advocacy groups to co-design practical approaches for communicating brain injury-related data with families in sensitive and timely ways that support informed choice and personalised care while considering the potential for anxiety or confusion. Such approaches might include improving access and signposting to trusted information resources, promoting awareness of antenatal screening options, co-designing antenatal discussion tools accessible and appropriate for the range of populations accessing maternity services, or piloting new ways of explaining how teams identify and respond to concerns or risks in specific services or to specific conditions. These approaches should be reviewed over time and informed by ongoing feedback from people with diverse backgrounds and needs, including those with lived experience of trauma or bereavement, and those who have navigated complex care needs following a diagnosis of brain injury.

Brain injury-related data should be shared in ways that recognise that how and when information is presented and contextualised can shape how it is received, particularly during an emotionally charged time like pregnancy. When national bodies or services share data, those data should be accompanied by clear context – for example, how teams identify and respond to risks and concerns and what safe care looks like – instead of delivering standalone statistics. Where appropriate, summaries of learning already implemented from past cases (from which data may have been derived), such as recurring contributory factors, or improvements since made, may support conversations between professionals and parents or others engaging with maternity services.

Sharing should be embedded into prenatal and antenatal care pathways, such as during midwife appointments or birth planning conversations. Professionals should be supported to answer questions and signpost people to trusted sources of information.
